# Deep-learning-derived glaucoma-related endophenotypes enable novel genome-wide genetic and functional discovery

**DOI:** 10.1101/2025.11.04.25339517

**Authors:** Liyin Chen, Yan Zhao, Saber Kazeminasab Hashemabad, Tobias Elze, Mohammad Eslami, Mengyu Wang, Janey L. Wiggs, Ayellet V. Segre, Nazlee Zebardast

## Abstract

The genetic architecture of primary open-angle glaucoma (POAG), a leading cause of irreversible blindness, remains largely unexplained due to the reliance of previous genome-wide association studies (GWAS) on imprecise phenotypes from electronic health records. Here, we overcome this with a disease-trained, task-transfer machine learning (ML) framework that learns glaucoma-related damage patterns from a large clinical repository of 8,323 glaucoma patients. We showed that ML optical coherence tomography (OCT)-derived endophenotypes trained on 18,985 OCT scans from these patients identified novel loci associated with POAG. By applying the derived endophenotypes to 47,908 UK Biobank participants, we performed GWAS in European, African, and Asian ancestral groups followed by cross-ancestry meta-analyses. In total, we identified 36 and 43 LD-independent GWAS loci that passed genome-wide significance in the EUR and cross-ancestry meta-analysis, respectively. About two thirds of the identified loci overlapped with previously reported POAG related associations, demonstrating the validity of our approach. Importantly, more than a third (21) of the loci were novel to glaucoma. Extensive functional analyses, including Bayesian colocalization analysis, gene-based association tests, Mendelian randomization, and single-cell enrichment analysis, converged on 11 high-confidence gene effectors, five of which are novel to glaucoma. These genes support Wnt-mediated outflow dysfunction and retinal ganglion cell vulnerability in POAG pathogenesis and are potential actionable drug targets. Our findings expanded POAG genetic associations, provided mechanistic insights at cell-type resolution, and proposed plausible putative causal genes. This study provides a powerful, generalizable ML-driven strategy for accelerating the discovery of disease mechanisms and therapeutic targets for complex diseases.

**One Sentence Summary:** A ML framework that bridged clinical eye scans with biobank genetics found 21 new genetic loci and 11 putative causal genes for glaucoma

## INTRODUCTION

Glaucoma is a leading cause of irreversible blindness worldwide, affecting over 80 million people worldwide and is projected to reach >110 million by 2040 (*1, 2*). Because current treatments primarily reduce intraocular pressure (IOP), deeper molecular insight is essential to shift glaucoma care from damage limitation to neuroprotection and prevention. As glaucoma is a highly heritable disease, especially its most common form, primary open angle glaucoma (POAG), numerous genome-wide association studies (GWAS) have been carried out for this disease over the last two decades (*3, 4, 4–11*). Despite these efforts, currently known POAG-associated loci only explain about 9.4% of the disease heritability (*5, 12*).

Accurate disease phenotyping is necessary for successful GWAS, and a limitation of prior glaucoma GWAS is their reliance on diagnostic codes and case-control definitions. These conventional phenotyping methods suffer from noise inherent to diagnostic codes in electronic health records (*13, 14*). By contrast, studies based on clinically examined cases and controls are often underpowered due to smaller sample sizes. Moreover, the case-control phenotyping approach fails to capture of full spectrum of disease heterogeneity. To address these limitations, intermediate quantitative endophenotypes are often used, because they are measured on a continuous scale, offering greater statistical power than dichotomous labels. Using a multi-trait analysis, prior work has demonstrated the utility of endophenotypes such as intraocular pressure (IOP) (*15*) and vertical cup-to-disk-ratio (vCDR) for genome-wide discovery for POAG (*16*). However, while these endophenotypes significantly improved GWAS power, they are indirect and imperfect surrogate for glaucomatous damage(*17, 18*).

An objective indicator of glaucomatous change include thinning of the retinal nerve fiber layer (RNFL) and the ganglion cell complex (GCC) (*19*) as measured by optical coherence tomography (OCT) scans, routinely used in clinical practice. Several studies have performed GWAS on average retinal layer thicknesses calculated from OCT scans (*20–22*). The association signals from these studies spanned both loci previously implicated in ocular or systemic disease and loci that modulate normal inter-individual variation in retinal morphology, demonstrating that retinal layer thicknesses capture functional and structural variations in the retina. However, the use of global average thicknesses, which compress information from 2-D thickness maps to a singular value, miss spatially heterogeneous patterns of damage that are typical of glaucomatous eyes. For example, Currant et al., 2021 attempted to use the average inner retinal layer thickness as an endophenotype for POAG but only replicated one known POAG loci.

With the recent advances in machine learning (ML) methods, nuanced endophenotypes from medical images, including OCT-derived retinal thickness maps, have been extracted for enhanced genome-wide association studies (*23–27*). Recent studies sought to enhance genome-wide discovery by performing GWAS on latent representations of retinal structural variability extracted from OCT-derived thickness maps (*28, 29*). While these studies extracted richer features beyond global average layer thicknesses, they fail to capture disease-specific variation when trained entirely on a largely healthy UKBB population. As a result, identified endophenotypes reflect general morphological variation of the human retina rather than disease-relevant changes.

To allow for disease-specific genome-wide discovery, here, we developed a task-transfer approach to improve quantitative phenotyping for POAG. We used unsupervised machine learning and a large clinical repository of OCT scans from 8,323 open-angle glaucoma (OAG) patients to capture clinically-derived glaucoma-related damage patterns that are not captured in largely healthy population-based cohorts. We subsequently mapped our trained ML endophenotypes onto the OCT scans from 47,908 participants in the UKBB, a large biobank where genotype data is available. This approach leverages the large number of OAG patients in our clinical cohort while bridging both the paucity of glaucoma cases in UKBB and the absence of genotypes in the clinical cohort, enabling well-powered, disease-specific genetic studies.

We identified 36 and 43 GWAS-specific LD independent loci with genome-wide significance from the EUR and cross-ancestry meta-analysis, respectively. Importantly, 21 loci have not been previously reported to be associated with glaucoma or well-established glaucoma-related traits. We further explored the functional significance of these previously unidentified loci by performing gene-based association tests, colocalization analysis with expression and splicing quantitative trait loci (eQTL and sQTL), mendelian randomization, and cell type enrichment analysis. By integrating multiple lines of genetic evidence, we highlight 11 gene associations, including 5 novel ones, for their functional relevance to glaucoma, which can be further investigated as clinically interpretable therapeutic leads. Overall, by uniting disease-focused deep representation learning with cross-ancestry genetics and functional single-cell omics, we validated and extended the putative causal variants and genes for POAG. Beyond ophthalmology, our study establishes a scalable, generalizable framework for ML-driven phenotyping of complex diseases and novel genome-wide discovery.

## RESULTS

### Subhead 1: Study Design

We previously developed an ML method to extract quantitative endophenotypes from GCC and RNFL thickness maps derived from macular OCT scans(*30*). Briefly, two deep learning models, an unsupervised autoencoder (AUTO) and a self-supervised momentum contrastive learning-based method (MOCO), were used to learn the latent representations of the GCC and the RNFL thickness maps. The latent representations then underwent dimensionality reduction and gaussian mixture modeling for clustering. This method allows for the identification of distinct retinal layer endophenotypes for subsequent genetic studies. We have previously shown the clinical correlations of these endophenotypes.

The study design of this work is illustrated in **Fig. 1**. Specifically, we trained the two models described above (MOCO and AUTO) using 18,985 macular OCT scans of 8,323 glaucoma patients (99% with open-angle glaucoma [OAG]) treated at Massachusetts Eye and Ear (MEE). The trained model was then mapped to 86,115 OCT scans from 35,739 European (EUR), 1,677 Asian (ASN), and 1,730 African (AFR) participants with genotype array data in the UKBB (1,074 [0.027%] were diagnosed with glaucoma by ICD code and self-reports). Twenty-one endophenotypes, 14 from the AUTO model and 7 from the MOCO model, were found in UKBB.

**Fig 1.**
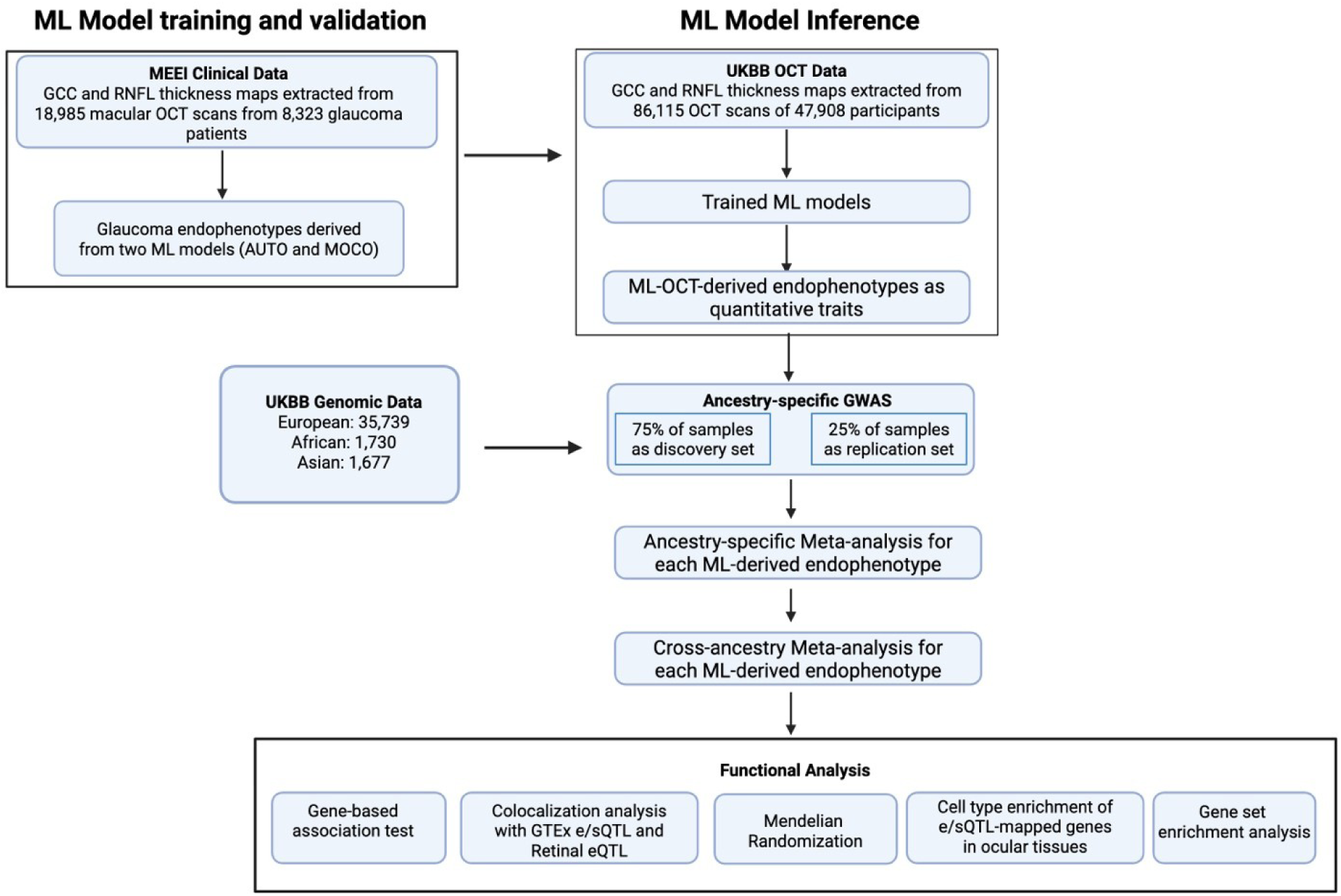
Overview of the study design. The flowchart illustrates the main steps of the study. First, ML models were trained on 18,985 OCT scans from 8,323 glaucoma patients at MEE to derive glaucoma-related endophenotypes. These trained models were then applied to 86,115 OCT scans from 47,908 UKBB participants to generate quantitative traits for genetic analysis. GWAS were performed separately on participants of European, African, and Asian ancestry using a 75% discovery and 25% replication set. The results were combined through ancestry-specific and subsequent cross-ancestry GWAS meta-analyses for each ML-OCT-derived endophenotype. Finally, a suite of functional analyses, including gene-based tests, colocalization, Mendelian randomization, and cell-type enrichment, was conducted to interpret the genetic findings and prioritize causal genes.

The genotype arrays in the UKBB were imputed and underwent quality controls, yielding an average of 8,590,260 common variants (minor allele frequency [MAF] >0.01) in each population for genetic association tests. Because of the lack of a separate cohort with both macular OCT scans and genotyping data for validation, to identify a high confidence set of variants, we divided the UKBB dataset from each population into a discovery and replication subset in a 3:1 ratio. GWAS was performed on each endophenotype and study subsets using a fixed effects linear regression model, adjusting for age at recruitment, sex, refractive error (measured as spherical equivalent) and top 10 genotype principal components (PCs), separately for each ancestral group, followed by meta-analyses of the discovery and replication subsets. A final cross-ancestry meta-analysis was conducted to enhance statistical power and identify shared genetic risk factors across populations. Post-GWAS functional analytical approaches were then applied to prioritize the genes and potential underlying causal mechanisms of the genetic findings for these endophenotypes.

### Subhead 2: GWAS and meta-analysis on ML-OCT-derived glaucoma endophenotypes

We performed GWAS of 21 total endophenotypes derived using our two ML methods in the EUR subsets. In the EUR discovery GWAS, 637 and 23 unique LD independent (r^2^ < 0.2) variants passed suggestive (P *≤* 1×10^-5^) and genome-wide significance (P *≤* 5×10^-8^), respectively (SI File 1). Of these, 63 and 14 unique variants passed nominal significance (P *≤* 0.05) and Bonferroni-correction in the replication GWAS, respectively. In the EUR meta-analysis of the discovery and replication subsets, 36 GWAS-specific LD-independent variants passed genome-wide significance (P *≤* 5×10^-8^) across the 21 endophenotypes (**Fig. 2a**, SI File 2). Among them, 17 passed the Bonferroni-corrected genome-wide significance adjusting for the number of endophenotypes (P *≤* 5×10^-8^ / 21). The effect sizes of these variants show high concordance (R^2^ = 0.93) between the EUR discovery and replication subsets (**Fig. 3a**).

**Fig. 2.**
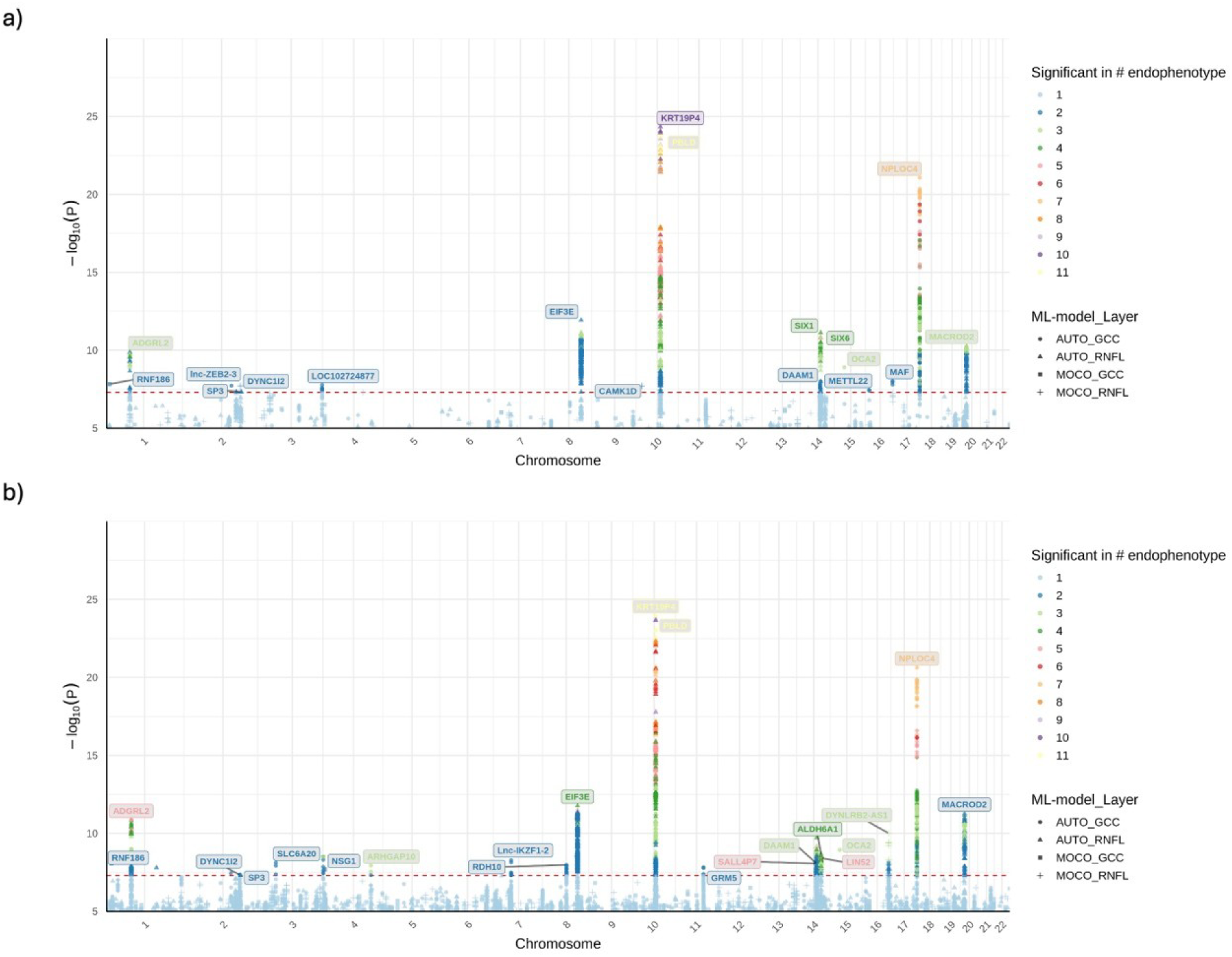
Overlayed Manhattan plots of GWAS meta-analyses for 21 ML-OCT-derived endophenotypes. Panel (a) displays the results from the EUR meta-analysis, while panel (b) shows the results from the cross-ancestry meta-analysis. In both plots, the y-axis represents the −log10(P-value) of association, and the x-axis shows the chromosomal position. The red dashed line indicates the threshold for genome-wide significance (P ≤ 5 × 10⁻⁸). Each point represents a genetic variant, with colors indicating the number of endophenotypes for which the variant reached significance. The shape of the points corresponds to the ML model and retinal layer for the most significant association at that locus. Nearest genes of the genome-wide significant loci are labeled.

**Fig 3.**
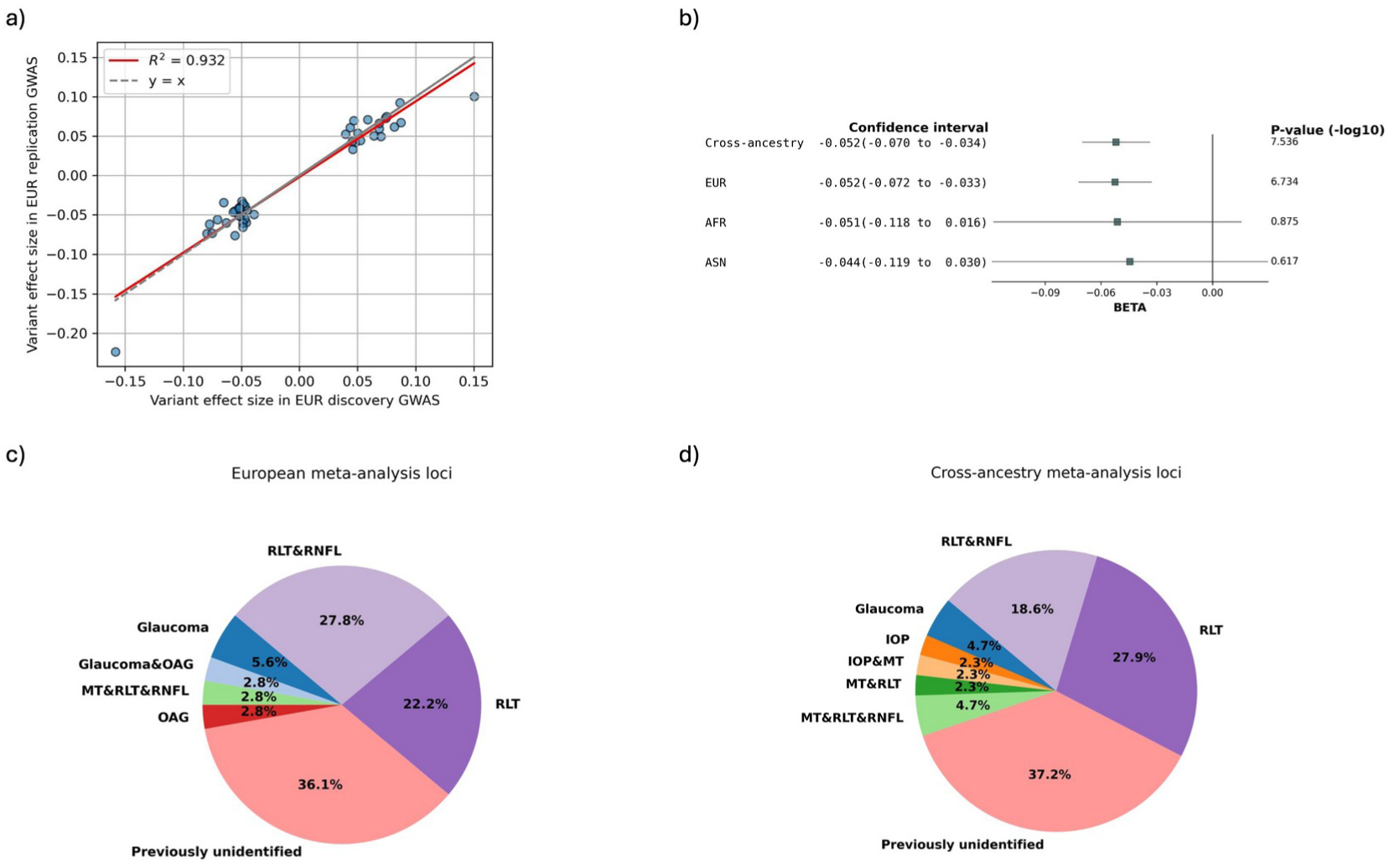
ML-OCT-derived endophenotypes replicated and expanded glaucoma genetic associations with consistent effect sizes across cohort subsets and ancestries. a) The effect sizes of significant loci in the EUR discovery GWAS stayed highly concordant in the replication GWAS. b) Representative forest plot to show that the effect sizes of genome-wide significant loci were relatively consistent across meta-analysis with different ancestries. The large standard errors in AFR and AMR were due to the small sample sizes. c) and d) Pie plots showing the proportions of EUR and cross-ancestry meta-analysis genome-wide significant loci that were previously reported in glaucoma, open-angle glaucoma (OAG), intraocular pressure (IOP), retinal nerve fiber layer thickness (RNFL), retinal layer thickness (RLT), and macular thickness (MT).

Due to the small sample sizes of the AFR and ASN groups, only 4 and 3 LD independent loci were identified, respectively for these population meta-analyses (SI Sheet 2). Nevertheless, the cross-ancestry meta-analysis identified 43 unique LD independent genome-wide significant loci across the 21 endophenotypes, with 20 passing the Bonferroni-corrected genome-wide significance (P *≤* 5×10^-8^ / 21) (Fig. 2b) (SI File 2). Of the 36 unique LD independent variants identified in the EUR meta-analysis, 49% (18) replicated in the cross-ancestry meta-analysis at genome-wide significance.

Effect-size heterogeneity (I²) for the genome-wide significant loci was minimal in both the EUR and cross-ancestry meta-analyses, indicating consistent genetic associations across GWAS subsets and across populations. We visualized the consistency of effect sizes across EUR and cross-ancestry meta-analyses using forest plots (**Fig. 3b**, SI File 3). All loci demonstrated highly consistent effect sizes between EUR and cross-ancestry analyses (Pearson correlation coefficient r = 0.998, P = 1.341 x 10^-52^). Most loci also exhibited consistency within AFR and ASN populations, although these effect estimates had larger standard errors and did not reach statistical significance, as expected due to the smaller sample sizes. None of the GWAS or meta-analyses showed any evidence of population structure inflation (λ=1.000-1.028) (SI File 4).

### Subhead 3: Genetic associations for glaucoma were replicated and expanded

We systematically compared the genome-wide significant loci identified in our EUR and cross-ancestry meta-analyses to previously reported associations with POAG and established glaucoma-related traits, including IOP, RNFL thickness and macular thickness. Of the LD independent genome-wide significant loci, 23 out of 36 (64%) in the EUR meta-analysis and 27 out of 43 (63%) in the cross-ancestry meta-analysis replicated known glaucoma-related genome-wide significant genetic associations (including LD proxies with r^2^ > 0.1) (**Fig. 3c**, **3d**) (SI File 5). The novel glaucoma loci found in this study were listed in Table 1. Multiple of them were overlapped between the EUR and the cross-ancestry meta-analyses, yielding 21 unique novel loci.

**Table 1.**
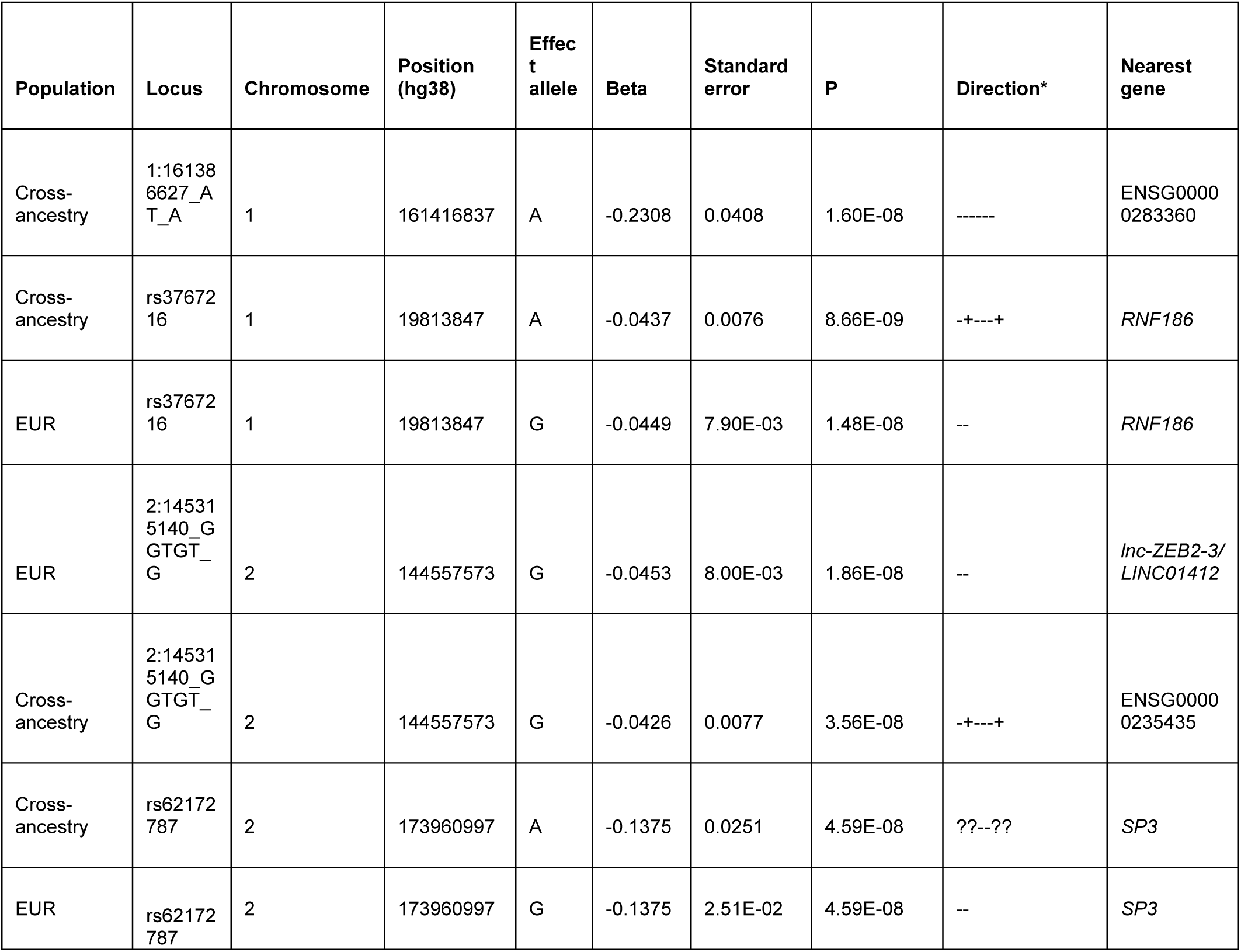
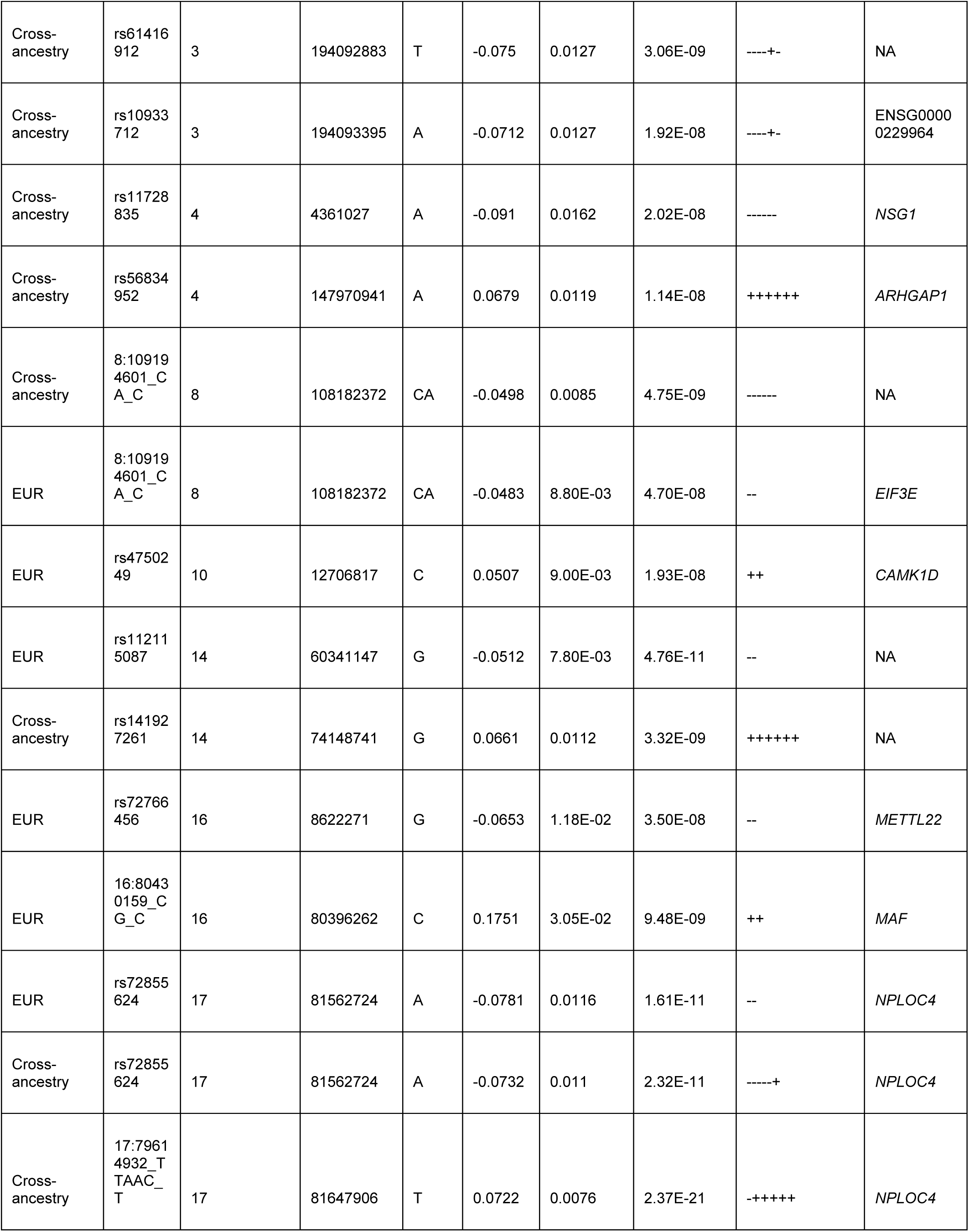
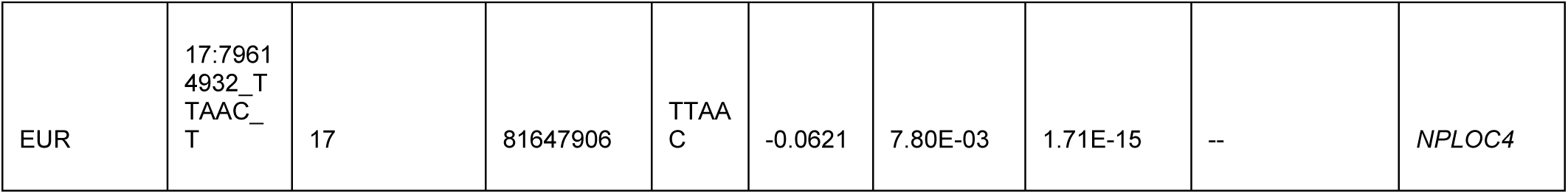
Novel loci from EUR and cross-ancestry ML-OCT endophenotype GWAS meta-analyses. The table lists the 21 novel, LD-independent loci that reached genome-wide significance (P ≤ 5 × 10⁻⁸) and have not been previously associated with glaucoma or its related traits. For each lead variant, the table details the population in which it was identified, its chromosomal position (genome build hg38), the effect allele, the effect size (beta) and standard error and association P-value from the corresponding meta-analysis, and the nearest mapped gene. *The direction of effect signs corresponds to the discovery and replication GWAS in the EUR meta-analysis, and discovery and replication for each of the populations in the following order: AFR, EUR and ASN.

### Subhead 4: Gene-based association test

We performed gene-based association analysis for each ML-OCT-derived endophenotype using MAGMA and identified 28 and 27 genes that passed the gene-based Bonferroni-corrected threshold (P < 0.05/18,658 genes tested) for both the EUR and cross-ancestry meta-analyses, respectively. Among them, 68% (19) of the genes significant in EUR were also significant in the cross-ancestry meta-analyses. Full gene-based tests results are in SI File 6.

### Subhead 5: Colocalization analysis of GWAS significant loci with *cis* eQTLs and *cis* sQTLs

To further propose putative causal genes that may underlie genome-wide significant loci for these endophenotypes through regulatory effects, we used e/sQTLs in brain, nerve, artery, and fibroblast tissues from the Genotype-Tissue Expression (GTEx) project (*31*), as well as peripheral retina eQTLs (*32*). We applied eCAVIAR, a Bayesian colocalization method, to the genome-wide significant loci from each of the endophenotypes’ meta-analyses that had at least one genome-wide significant hit. e/sQTLs from the selected tissues that overlapped each GWAS locus LD interval were included for colocalization.

We found that 72% (26/36) of EUR GWAS loci and 40% (17/42) of cross-ancestry GWAS loci identified in our study significantly colocalized with at least one eQTL and/or sQTL (colocalization posterior probability [CLPP] >0.01). The retinal eQTLs that significantly colocalized largely overlapped with significant e/sQTLs identified in GTEx tissues. In total, 25 genes from the EUR meta-analysis and 24 genes from the cross-ancestry meta-analysis showed significant colocalization with the identified genome-wide significant loci (SI File 7). Of these, 95% (20/25) and 79% (19/24) of the colocalizing genes in EUR and cross-ancestry meta-analysis were protein-coding, respectively.

**Figure 4** and Fig. S1 summarizes the e/sGenes that colocalized significantly with at least one of the genome-wide significant loci from the cross-ancestry and EUR meta-analysis, respectively. Several e/sGenes, including *EIF3E, PCNX4, PBLD, TSPAN10,* and *DNAJC12*, colocalized with the novel loci in this study. In addition, 36% (10) and 30% (8) of the significant genes from the EUR and cross-ancestry gene-based tests overlapped with the significant e/sGenes identified through the colocalization analysis, respectively.

**Fig. 4.**
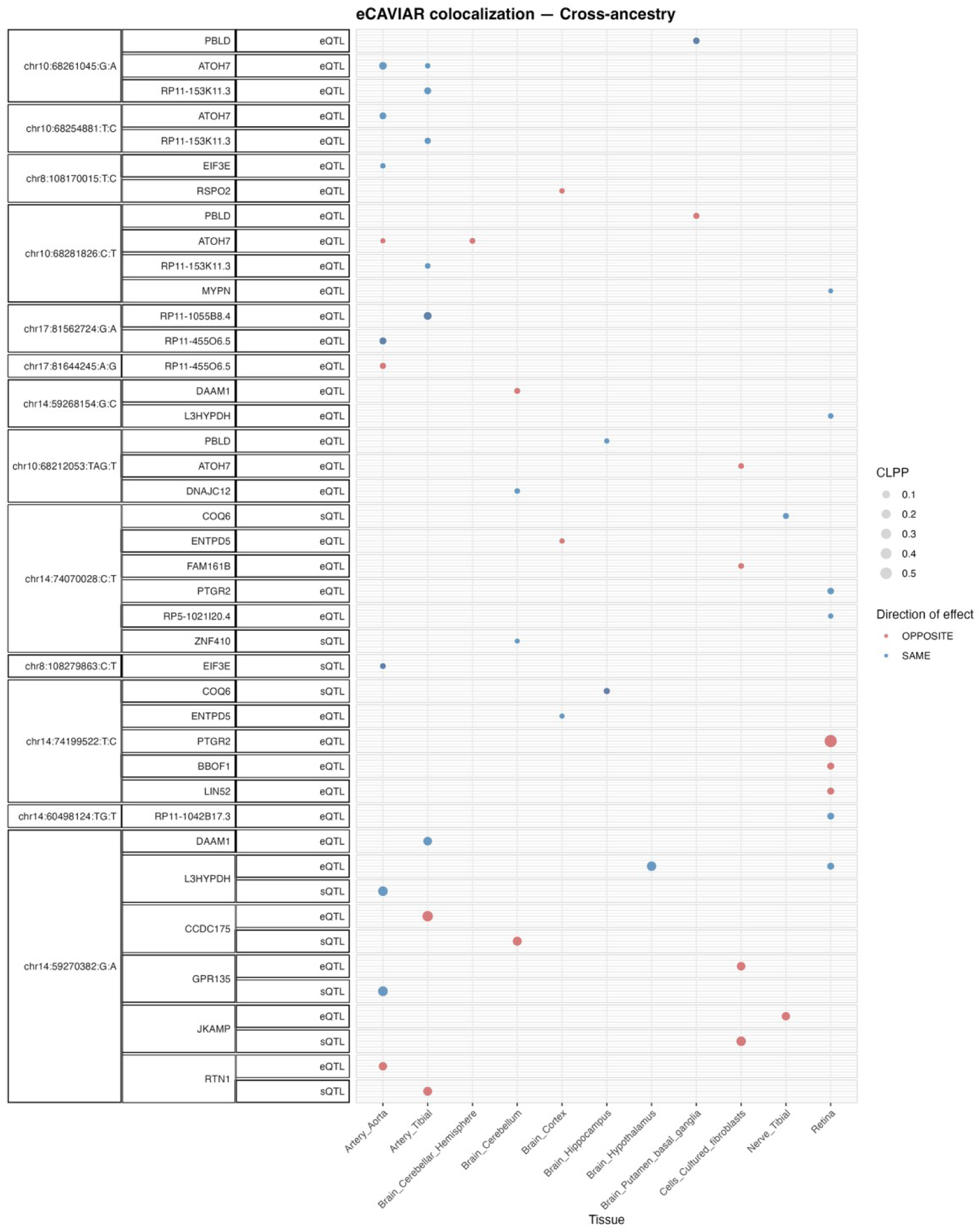
Colocalizing e/sQTLs in GTEx tissues and retina with genome-wide significant loci in the cross-ancestry meta-analyses. Shown are genes with at least one significant colocalization (CLPP > 0.01) between e/sQTLs tested across selected GTEx tissues and peripheral retina and genome-wide significant loci identified in the cross-ancestry meta-analysis. Bubble size represents the maximum colocalization posterior probability among all e/sVariants tested for each gene, QTL type, and tissue combination. Points are color-coded according to the relative direction of effect between the QTL and the GWAS meta-analysis.

### Subhead 6: Cell-type enrichment analysis

To connect the identified genes to involved cell types and disease mechanisms, we applied ECLIPSER (*33, 34*) to test whether the expression of the identified colocalizing e/sGenes was enriched in specific cell types in key eye tissues relevant to the pathophysiology of glaucoma, the outflow pathways, retina, and optic nerve head and surrounding tissues. Consistent with Hamel *et al.* (2024) (*35*), we observed nominal enrichment (P<0.05, FDR<0.2) of RNFL endophenotype mapped genes in fibroblasts in both the unconventional (ciliary body and iris) and conventional (trabecular meshwork/Schlemm’s canal) outflow pathways in the anterior segment, and fibroblasts and astrocytes in the optic nerve head (Table 2, SI File 8, **Fig. 5a**, Fig. S2, and Fig. S3). These recurring cell-type signals underscore the central importance of outflow pathway fibroblasts, retinal support cells, and ONH macroglia in glaucoma pathogenesis and validate the robustness of our analysis.

**Figure 5.**
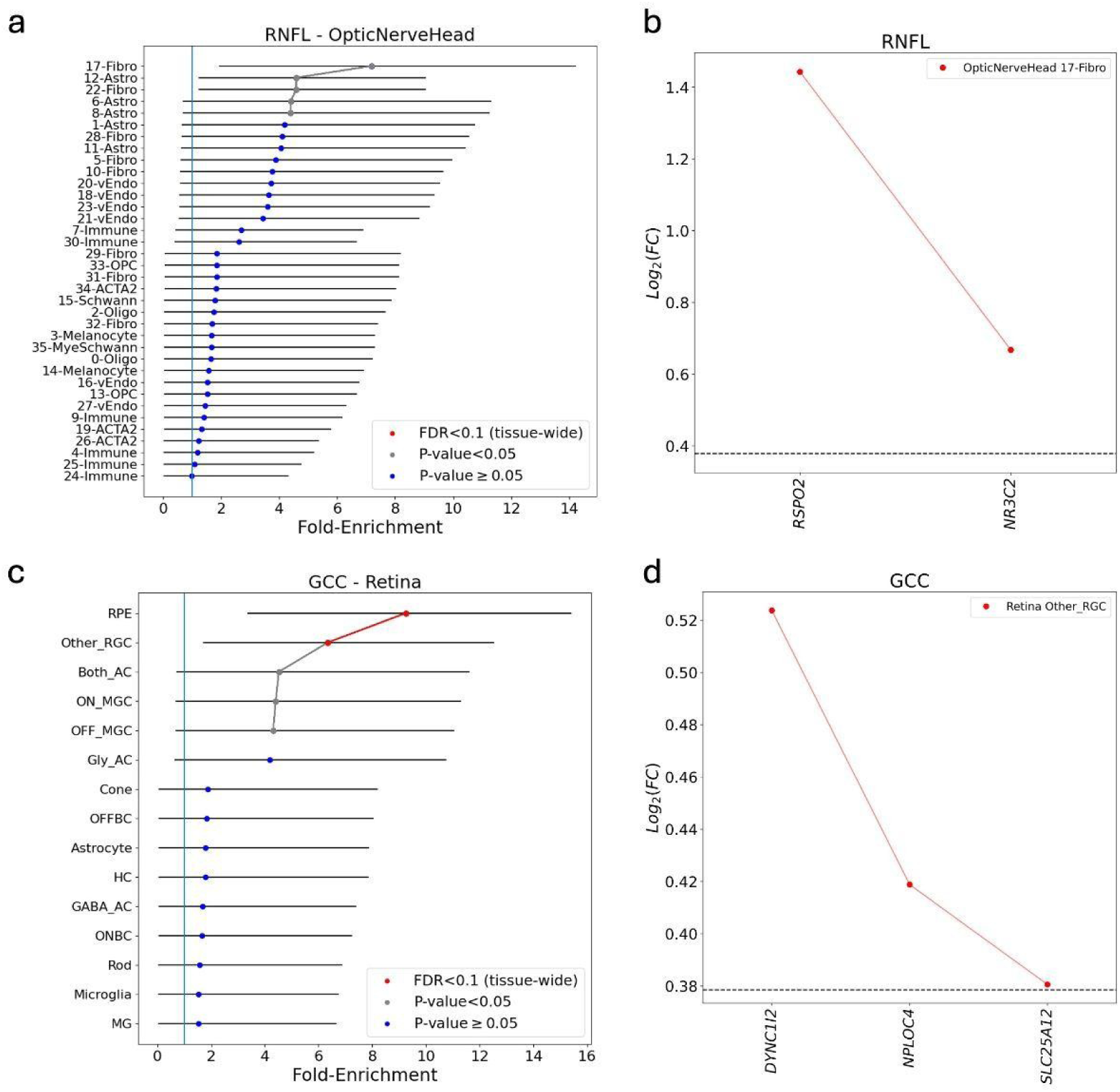
Cell type enrichment analysis of the cross-ancestry meta-analysis in the optic nerve head and retina. Panels (a) and (c) show the fold-enrichment of cell type-specific expression for genes mapped from the cross-ancestry GWAS meta-analysis loci in the optic nerve head for RNFL-derived endophenotypes and retina for GCC-derived endophenotypes, respectively. Cell types are ranked by their enrichment score (x-axis). Points are colored by statistical significance: red indicates tissue-wide significance (FDR < 0.1), while grey indicates nominal significance (P < 0.05). Panels (b) and (d) highlight the specific genes driving the most significant enrichment signals from the corresponding plots on the left. Panel (b) shows that *RSPO2* is the primary driver of the enrichment in optic nerve head fibroblasts. Panel (d) shows that *DYNC1I2*, *NPLOC4*, and *SLC25A12* collectively drive the enrichment in RGC clusters. The horizontal dashed line represents log2(Fold-change) of 0.375 (FC = 1.3) and FDR < 0.1 used as the cell type-specificity enrichment cutoff. FC, fold-change of gene expression in given cell type relative to all other cells in the given tissue.

**Table 2.** Summary of the ocular cell types showing significant enrichment of expression of e/sQTL-mapped genes to endophenotype loci. An asterisk (*) denotes cell types that passed the stringent tissue-wide significance threshold (BH FDR<0.1), while all other listed cell types passed nominal significance (P<0.05). Cell types are listed in order of significance in GCC and RNFL endophenotypes. Cell type descriptions are available in Tables S19 (cross-ancestry meta-analyses) and S20 (EUR meta-analyses).

| Tissue | Cross-ancestry,<br>GCC | Cross-ancestry,<br>RNFL | EUR, GCC | EUR, RNFL |
| --- | --- | --- | --- | --- |
| Anterior segment<br>(trabecular<br>meshwork/Schlemm's<br>canal, iris, central<br>cornea, corneoscleral<br>wedge, lens) | Iris_sphincter*,<br>CB_NPCE,<br>goblet, lens_fiber,<br>Uveal_Melanocyte | goblet, CB_NPCE,<br>iris_fibro,<br>ciliary_fibro, K_fibro,<br>iris_RPE, TM_fibro,<br>K_Endo | CB_NPCE,<br>Lens_Fiber,<br>Uveal_Melanocyte,<br>CB_PCE,<br>Iris_PPE,<br>Conj_Melanocyte,<br>Iris_APE,<br>Lens_EarlyFiber,<br>Iris_Sphincter,<br>Pericyte1 | CB_NPCE,<br>Iris_Fibro,<br>Ciliary_Fibro,<br>Iris_PPE,<br>TM_Fibro, Goblet |
| Retina | RPE*, Both_AC,<br>ON_MGC,<br>OFF_MGC | Astrocyte | RPE*, Both_AC,<br>Other_RGC, Cone |  |
| Optic nerve head,<br>optic nerve and<br>surrounding posterior<br>tissues | Fibroblasts,<br>melanocytes,<br>immune cells,<br>6_Astro, 8_Astro | Fibroblasts,<br>astrocytes, 6_Astro,<br>8_Astro | 3-Melanocyte*,<br>14-Melanocyte* | 17-Fibro, 28-Fibro,<br>5-Fibro, 10-Fibro,<br>22-Fibro, 22-Astro |
| Macula | RPE*, | H2 | RPE*, H2 |  |

Importantly, our analysis highlighted several enriched cell types and lead genes that were previously unreported but may mediate glaucoma risk. Nominal enrichment was observed in fibroblasts of several anterior and posterior segment cell types in the EUR and cross-ancestry meta-analyses, which were all driven by *RSPO2* (**Fig. 5a**, **5b**). Notably, *RSPO2* appeared repeatedly in our functional analyses. One of the most significant cell type enrichments found for GCC endophenotype genes (tissue-wide Benjamini-Hochberg adj. P<0.05) was in the retinal ganglion cell (RGC) cluster of all RGCs other than ON-MGC and OFF-MGC cells in the retina, that was primarily driven by *DYNC1I2*, *NPLOC4*, and *SLC25A12* (**Fig 5c**, **5d**). Additional cell types showing tissue-wide significance for the GCC endophenotype loci included optic nerve head melanocytes, retinal/macular RPE cells (Fig. 5c) and iris sphincter muscle cells. The pigmented cell type enrichments were driven by genes (*OCA2, NPLOC4 and TSPAN10*) associated with myopia(*36*), a risk factor for POAG.

### Subhead 7: Mendelian randomization of putative causal genes

To provide additional support for a causal relationship between e/sQTLs and our glaucoma endophenotype associations, we applied Generalized Summary-data-based Mendelian Randomization (GSMR) to the 12 genes that show statistical significance in at least two of the three functional analyses we applied to the endophenotype GWAS: colocalization analysis, gene-based association testing, and cell-type enrichment analysis (Table 3). After removing variants with horizontal pleiotropy, 92% (11) of genes showed evidence of causality using a Bonferroni-corrected threshold (P < 0.05/13) accounting for the number of genes tested. Complete MR results are available in the SI File 9.

**Table 3.**
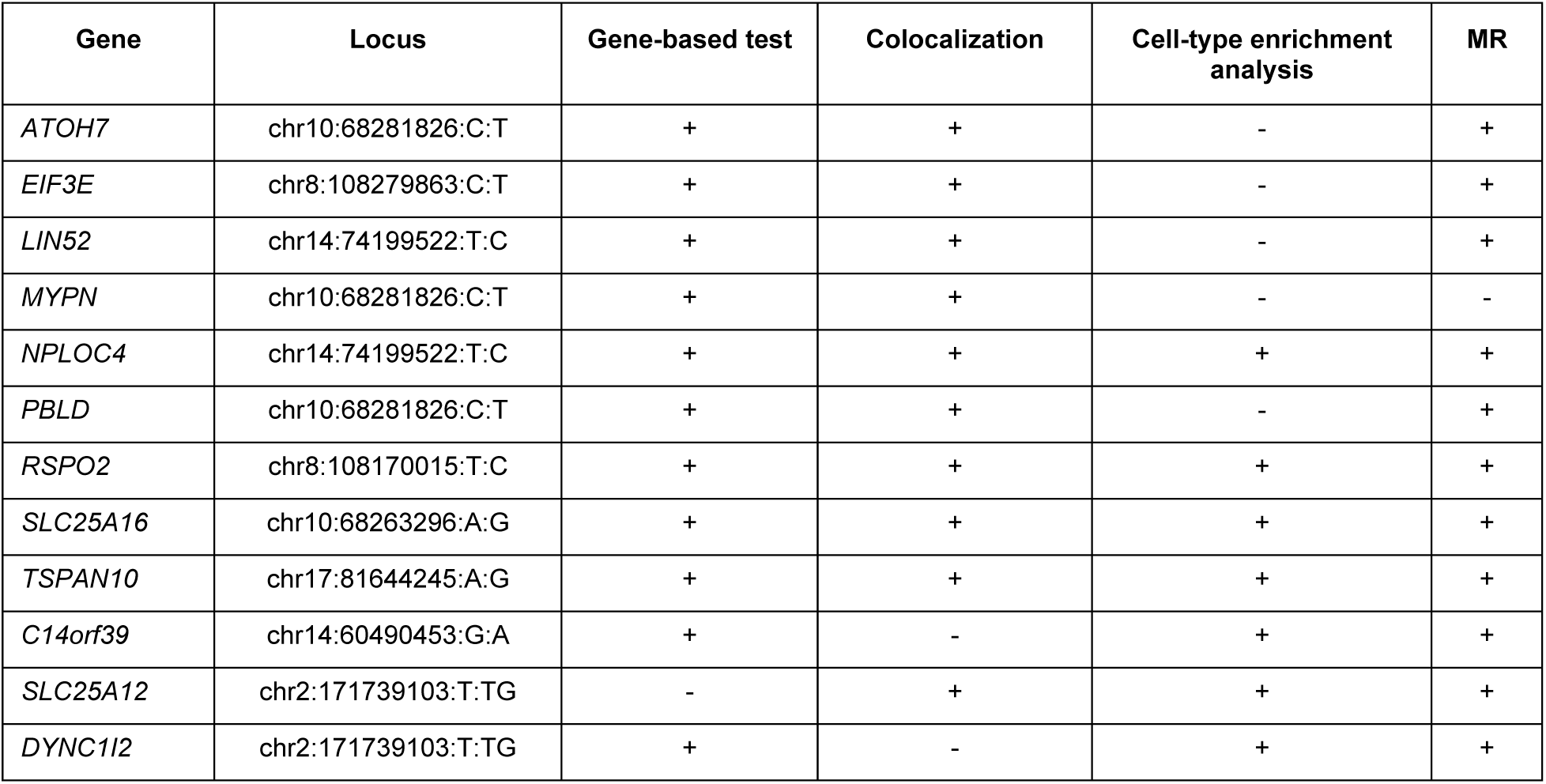
Summary of functional evidence for prioritized genes. The table highlights genes that were statistically significant in at least two of the primary functional analyses performed. A "+" indicates that a gene passed the significance threshold for the corresponding analysis; A "-" indicates the gene did not meet the significance threshold for that analysis. MR, Mendelian randomization.

### Subhead 8: Gene-set enrichment analysis

Gene-set enrichment analysis of the endophenotype GWAS using MAGMA recapitulated known POAG biology while sharpening the axes involved. In both the EUR and cross-ancestry meta-analyses, we observed robust evidence for lipid/cholesterol regulation and innate inflammatory programs, highlighted by enrichment of SREBF1A targets (P = 8.31 x 10^-3^, SI File 10), response to oxidized phospholipids (P = 1.22 x 10^-4^), IL-17/IL-22 signaling (P = 2.07 x 10^-2^) and suppression of phagosome maturation (P = 2.80 x 10^-5^). Vascular and hypoxic responses were prominent (VEGFR1/2 signaling, P = 1.54 x 10^-5^; HIF1/2 targets, P = 2.66 x 10^-3^), as well as developmental/ECM-remodeling cascades relevant to aqueous outflow and retinal support tissues (WNT signaling and Notch activation). We also observed the calcium signaling pathway enriched in both analyses (P = 1.98 x 10^-4^). Our results reinforced and extended the pathway landscape reported by recent large POAG GWAS(*12, 35*), providing support for altered lipid handling, para-inflammation, vascular/hypoxic stress, and tissue-remodeling signaling as biological processes influencing POAG.

## DISCUSSION

Our study shows that disease-focused, deep-learning phenotypes extracted from macular OCT can expand the genetic understanding of POAG. Our European and cross-ancestry meta-analyses uncovered 36 and 43 genome-wide significant loci, respectively. Among these loci, roughly 65% were previously reported to be associated with OAG or at least one well-established glaucoma-related trait, such as IOP and retinal layer thickness. The concordance of our GWAS hits with established glaucoma loci suggest that our ML-OCT-derived endophenotypes are disease specific and able to capture contributions of glaucomatous structural damage. More importantly, however, we identified 21 novel genome-wide significant loci, where several of the mapped genes are highly expressed in relevant eye tissues, such as ONH and retina. Biological relevance of the identified loci is further supported by multiple lines of evidence.

Using systematic functional analyses, we advance from association to higher-resolution gene and cell-type inference, providing a basis for mechanistic hypotheses that can be followed up for POAG. Bayesian colocalization with multiple GTEx tissues that contain relevant cell types plus retinal eQTLs linked 72 % of European association signals to likely regulatory gene/s in tissues with eye-relevant cell types. MAGMA gene-based tests uncovered 28 significant genes from the European meta-analysis, 68 % of which overlapped with those from the cross-ancestry analysis. Importantly, 36 % of MAGMA hits in EUR cohort overlapped colocalizing e/sGenes, providing further statistical support. Cell-type specific enrichment analysis connected these genes to the cellular architecture of the eye, where pigmented cells in the retina and ONH, ONH and outflow pathway fibroblasts, and ONH astrocytes that emerged as the most over-represented populations, echoing the cell-types produced by Hamel *et al.* (2024). The enrichment in outflow pathway cells suggested that elevated IOP also plays a role in affecting glaucomatous damage in the retina. Not previously observed with POAG GWAS loci was enrichment in non-midget ganglion cell RGCs driven by *NPLOC4, SLC25A12*, and *DYNC1I2*. Mendelian randomization further strengthened the role of the identified genes in glaucoma pathogenesis. This layered approach prioritized 11 putative causal genes (*EIF3E, LIN52, NPLOC4, PBLD, RSPO2, SLC25A16, TSPAN10, DYNC1I2, SLC25A12, ATOH7*, and *C14orf39)*.

Among the prioritized genes, some are known to be associated with POAG related traits, while others are novel. Specifically, *ATOH7* and *C14orf39* are genes that have been previously proposed to potentially contribute to POAG risk (*37, 38*). *SLC25A16,* and *PBLD* are genes near *ATOH7* and have been reported to be associated with VDCR. *TSPAN10* and *NPLOC4* have been reported to be associated with ocular traits like macular thickness and optic disc parameters. Importantly, five genes (*RSPO2, EIF3E, LIN52, DYNC1I2, SLC25A12*) that showed strong supportive causal evidence in our studies have not been reported previously. For example, *EIF3E*, a gene from one of our novel loci, highly significant in the gene-based association test (P = 4.52 × 10⁻¹³), showed a significant colocalization signal with multiple brain and artery tissues, and had elevated expression across several outflow pathway, ONH and retinal cell types (Fig. S4). As another example, *RSPO2* was significant in gene-based association tests (P = 2.13 × 10^-9^) and colocalized with eQTLs in brain cortex. Single-cell expression in ocular cell types showed elevated *RSPO2* expression in the fibroblasts of ONH and multiple cell types in the anterior segment, with the highest expression levels in fibroblasts in the trabecular meshwork, iris and ciliary body. Cell-type enrichment analysis further supported *RSPO2* as the main driver of fibroblast enrichment in these regions.

The genes and ocular cell types identified in this study point to two connected biologic mechanisms of POAG risk. *RSPO2* is a potentiator of canonical Wnt signaling (*39*); its high expression in anterior-segment and ONH fibroblasts suggest a role in extracellular-matrix remodeling and aqueous outflow regulation that potentially leads to elevated IOP. On the neuronal side, *EIF3E*, an initiator for protein synthesis and DNA damage repair (*40*), shows strong gene-based and colocalization support with broad retinal expressions, implicating a potential role in RGC resilience. *DYNC1I2* underlies enrichment in an RGC subtype clusters and encodes a dynein-1 intermediate chain; dynein-dependent axonal transport is known to fail early in experimental glaucoma (*41*), potentially linking this locus to RGC transport vulnerability. *SLC25A12*, a mitochondrial aspartate–glutamate carrier, contributes to the same RGC enrichment and supports neuronal energy metabolism. This data supports a model where Wnt-tuned fibroblast programs in the ONH and outflow pathway modulate IOP and biomechanical stress, which then interface with RGC-intrinsic capacities for protein synthesis/repair, axonal transport, and mitochondrial bioenergetics to determine susceptibility to glaucomatous injury.

While these novel genes do not have established therapeutic agents, publicly available data suggests they represent highly tractable targets for future drug discovery programs(*42*). For instance, *RSPO2*, *EIF3E*, and *DYNC1I2* possess known protein-ligand structures that can guide small molecule development. Specifically, the existence of a co-crystal structure of *RSPO2* with a small molecule ligand provides a direct blueprint for structure-based drug design. Furthermore, as *RSPO2* is a secreted potentiator of canonical Wnt signaling highly expressed in anterior-segment fibroblasts, developing a neutralizing antibody represents a highly feasible and specific therapeutic strategy. These results demonstrated that our genetic findings not only contributed to the understanding of POAG biology, but also proposed high-confidence starting points for functional validations with patient-derived cells and animal models, ultimately paving the way for novel IOP-lowering and neuroprotective treatments for POAG.

There are several limitations in this work. While our ML models were trained on a large group of patients with POAG related ICD codes, inherent coding errors may introduce some noise. While not feasible at this scale, focusing on patients with confirmed POAG, as verified by chart review, may have resulted in more specific POAG endophenotypes. Nevertheless, we expect the size of our dataset largely overcomes this limitation. Additionally, due to the lack of an external cohort with available OCT and genotype data, GWAS replication was performed by a 75:25 split of the UK Biobank samples, a less stringent approach than true external replication, but represents the most rigorous validation currently feasible at this scale. Importantly, the strong concordance of effect sizes between our discovery and replication sets, combined with extensive validation from multiple downstream functional analyses, provides high confidence in our findings and establishes a powerful phenotyping framework for future replication efforts. Regarding the association analyses, although the first ten genotype principal components were included as covariates, subtle population substructure may persist and inflate association signals. Finally, the generalizability of UK Biobank results is limited due to its European-centric sample. Confirmation of the smaller but directionally aligned effects seen in our African and Asian groups is needed using larger non-European OCT cohorts.

In conclusion, by integrating disease-trained ML-OCT-derived endophenotypes with systematic functional genomics, we have successfully expanded the number of glaucoma associated loci, linked them to plausible causal genes, and mapped them to specific eye cell types. Beyond advancing our understanding of glaucoma’s genetic mechanisms, this study demonstrates the effectiveness of enhancing disease phenotyping by transfer learning from a clinical dataset to a population cohort, a methodology applicable to the investigation of other complex diseases.

## MATERIALS AND METHODS

### MEE cohort definition

OCT scans from glaucoma patients seen in MEE between October 1, 2020 to June 1, 2023 were used for model training. Glaucoma Individuals were defined based on the International Classification of Diseases (ICD) code in the electronic health record (i.e ICD9: 365.x, ICD10: H40.x). Specifically, 99% (8207) of these patients contain at least one of the OAG ICD codes. The other 1% are patients with other types of glaucoma, including angle-closure glaucoma and secondary glaucoma.

### Quality control of UKBB genotype data

Genotype array quality control steps were applied with PLINK v1.9 and PLINK v2.0 (https://www.cog-genomics.org/plink/) on directly genotyped and imputed variants of 488,395 UKBB samples. The hard call sample quality control (QC) was conducted to remove withdrawn samples (N=107), to exclude samples that are not in impute data (N=969), to exclude samples with no sample QC data (N=112), to remove ambiguous sex (N=1930), to remove heterozygosity outliers by using 5 standard deviations from the mean heterozygosity rate within each inferred ancestry subpopulation (N=978) and to remove related individuals with high cryptic relatedness (> 0.1875 Pi-hat) (N=35,113). To infer ancestry, Principal Component Analysis (PCA) was first applied to linkage disequilibrium (LD)-pruned (r2<0.2 in 200kb windows) genetic markers with MAF > 5%. The SHARDS algorithm selected the first 5 important PCs and then implemented k-nearest neighbors classification to predict the ancestral background of participants using labels from the 1000 Genomes Project Phase 3 reference panel. Participants were then categorized based on primary genetic ancestry. In terms of the relative removal algorithm, the graph theory was also applied to keep the maximal independent set. The application of maximal independent set contributed to keep the as many samples as possible. Concurrently, another relative removal algorithm was also conducted to keep highest ratio of the number of cases versus the number of controls to improve the power.

Regarding the imputed variant QC, the pipeline iteratively examines variants, INFO score, allele frequencies and tests of Hardy-Weinberg equilibrium. More specifically, the missingness rate per sample using posterior probability>0.9 for genotype calling was inspected. A small missingness rate of autosomes was observed and no significant difference was observed between male and female. Monomorphic sites (M=552,024) were removed if only one allele occurs at a site or locus in the population. We further filtered out the variants with indels length>50 (M=2,312) and subsequently, the variants with INFO score<0.6 (M=45,927,754) were excluded. The variants with allele frequency<0.1% for each ancestry subpopulation (EUR: M=35,850,305; AMR: M=28,792,905; AFR: M=24,954,341; SAS: M=35,437,737; EAS: M=37,236,001) were further excluded. Then the Hardy-Weinberg equilibrium was calculated and inspected for each ancestry subpopulation. The mid p-value was utilized in exact tests for Hardy-Weinberg equilibrium analyses. Therefore, a total of 227,173 variants in EUR, 98 variants in AMR, 32,317 variants in AFR, 38,173 variants in SAS, and 3,711 variants in EAS of autosomes failed Hardy-Weinberg and were removed. To be conservative, a variant was kept if it passed the Hardy-Weinberg for all ancestry subpopulations. Another MAF filter was applied to keep the common variants with MAF>1% for further analyses. After QC, the number of variants in EUR, AFR, and ASN are 9,329,555, 8,477,280, and 7,963,945, respectively.

### GWAS and meta-analysis

Macular GCC and RNFL endophenotypes were derived from 18,985 OCT scans of 8,323 glaucoma patients treated at MEE. These endophenotypes were mapped to OCT scans from the UK Biobank, including 35,739 participants of European ancestry, 1,730 of African ancestry, and 1,677 of Asian ancestry. For patients with scans from both eyes and multiple visits, we prioritized the right eye. If no scans were available for the right eye, the left eye was used instead. When multiple scans existed for the same eye, the most recent scan was used as the endo phenotype.

Genome-wide association studies (GWAS) were conducted on a total of 21 endo phenotypes, including 7 AUTO GCC phenotypes, 2 MOCO GCC phenotypes, 7 AUTO RNFL phenotypes, and 5 MOCO RNFL phenotypes. Each endophenotype was inverse rank normalized prior to performing GWAS testing. GWAS was performed separately for each ancestry group (AFR, Asian, and EUR). Within each ancestry, samples were randomly divided into discovery (75%) and replication (25%) sets, with matched age distributions and female/male proportions across the two subsets. Analyses were performed using PLINK v2.0, adjusting for age, sex, the first 10 principal components (PCs), and refractive error. PCs were calculated separately within each ancestry group. For meta-analysis, we used METAL (version 2011-03-25), applying a fixed-effects model based on inverse-variance weighting, under the assumption that all samples came from the same biobank. Heterogeneity statistics were computed, and beta coefficients and standard errors were used as inputs. Meta-analyses were firstly conducted within each ancestry, followed by a trans-ancestry meta-analysis.

LD clumping was performed with PLINK v2.0 to identify independent loci in each meta-analysis. LD calculations were based on population-specific samples in the UKBB. For genome-wide significant signals, the LD threshold r^2^ was set to be 0.2 and the LD radius was set to be 500kb for LD clumping.

None of the GWAS or meta-analyses showed any evidence of population structure inflation, with the genomic lambda values around 1. Effect-size heterogeneity (I²) for the genome-wide significant loci was minimal in both the EUR and cross-ancestry meta-analyses, with most loci showing I² values near zero. Additionally, heterogeneity p-values for all tested loci were greater than 0.05, indicating consistent effects across populations (SI File 2).

### Comparison with previously reported associations

We compared our identified independent genome-wide significant loci to genome-wide significant variants previously reported to be associated with glaucoma and glaucoma-related traits, which are retinal layer thickness, IOP, CDR. The set of variants used for comparison were downloaded from GWAS Catalog (MONDO_0005041, EFO_0004190, OBA_2050110, EFO_0004695) and from three POAG GWAS studies (*35, 43, 44*). To ensure that our classification of novel vs previously reported loci is LD-aware, we included LD proxies (r^2^ > 0.1, UKBB as reference panel) of the identified loci in this comparison.

### Colocalization analysis with expression and splicing QTL datasets

We conducted a Bayesian colocalization analysis using eCAVIAR (https://github.com/fhormoz/caviar) to pinpoint candidate genes and regulatory elements (e/sQTLs) potentially involved in how glaucoma-linked genetic variations exert their effects. This method estimates the probability that overlapping GWAS and e/sQTL signals share the same causal variant or haplotype. This method considers local LD and the presence of multiple causal variants (allelic heterogeneity).

We used cis-expression quantitative trait loci (cis-eQTLs) and cis-splicing quantitative trait loci (cis-sQTLs) data from multiple tissues in the GTEx project (v8 release)(*45*) and peripheral retina eQTLs from the EyeGEx dataset(*46*). The GTEx data was based on GENCODE v26 annotations, while EyeGEx data used GENCODE v25. Significant e/sQTLs (FDR < 0.05), gene expression levels, and intron excision ratios (LeafCutter values) were downloaded from the GTEx portal (https://gtexportal.org/home/datasets) for GTEx and EyeGEx. These summary statistics for all tested variant-gene pairs were then used for colocalization analysis.

Colocalization was performed on the genome-wide significant LD-independent loci for each GWAS meta-analysis. Every GWAS locus was assessed for colocalization with all eQTLs and sQTLs containing at least five e/sVariants (FDR < 0.05) located within the corresponding GWAS LD interval. For each locus, LD windows were defined by selecting variants in LD (r² > 0.1) with the lead GWAS variant, extending 50 kb upstream and downstream. For loci derived from European populations, LD calculations were based on European samples from the 1000 Genomes Project (*47*), while for the cross-ancestry loci, LD was computed using the European, African, and Asian samples in 1000 Genomes, using PLINK v1.9. When the lead variant was missing from the 1000 Genomes dataset, a high LD proxy (r² > 0.8) or the nearest variant was used. Z-scores for GWAS and e/sQTL summary statistics were computed as the effect size divided by its standard error for each variant. For retina-specific eQTLs, Z-scores were derived from association p-values, assuming a chi-square distribution with 1 degree of freedom. We tested all variants with MAF > 1% that were common to the GWAS and e/sQTL datasets. Variants in GWAS datasets were aligned to the alternative allele (ALT) reported in GTEx and EyeGEx datasets. Only eQTLs were analyzed for retina-specific data, while both eQTLs and sQTLs were analyzed for GTEx tissues. A locus was considered to exhibit significant colocalization when the colocalization posterior probability (CLPP) exceeded 0.01, following eCAVIAR’s recommendations. To reduce false-positive findings, we excluded any variant– gene–tissue–trait combinations in which the colocalized e/sVariant had a GWAS p-value > 0.05, an e/sQTL p-value > 1 × 10⁻⁴, or failed to meet the FDR < 0.05 criterion (i.e., labeled “FALSE” in the column ‘QC_name_QTL_FDR05_P1E04_GWAS_P5E02’ of SI Sheet 6).

### Gene-based tests and gene set enrichment analyses

We conducted gene-based association tests and gene set enrichment analyses with MAGMA (v1.10, https://ctg.cncr.nl/software/magma). In this gene-based analysis, genetic variants were mapped to 18,685 genes based on distance (+/-100kb) to compute a gene-level z-score, which was then tested for association with the ML-derived endophenotypes adjusting for cofounding factors. The derived gene-based P values were adjusted using the Bonferroni method to account for multiple testing (P < 0.05/18,685 = 2.68 × 10−6). In the pathway analysis, a list of 17,023 curated gene sets from MSigDB (https://www.gsea-msigdb.org/gsea/msigdb/) human database were used, and the P values of pathway analysis were adjusted by applying a Bonferroni correction.

### Mendelian randomization

For each gene identified by our colocalization or our gene-based analysis, we performed summary-data Mendelian randomization using Generalized Summary-data-based Mendelian Randomization (GSMR, https://yanglab.westlake.edu.cn/software/gsmr/) to test for a putative causal effect of cis-regulated expression on the endophenotypes. For each target gene, we first extracted independent cis-eQTL instruments (defined as SNPs within ±1 Mb of the transcription start site with P < 5 × 10⁻⁸) from the GTEx and retina eQTL summary statistics. Instruments were LD-pruned at r² < 0.05 in 1 Mb windows using a European-ancestry reference panel (n = 5,000 unrelated UK Biobank participants). To guard against horizontal pleiotropy, we applied the HEIDI-outlier test implemented in GSMR with a PHEIDI threshold of 0.01, removing SNPs inconsistent with a single causal pathway. GSMR then estimated the causal effect of gene expression on the endophenotype with a generalized least-squares framework that accounts for sampling variance and residual LD among instruments. Effect estimates and their standard errors were computed under the default GSMR settings.

### Ocular single-cell expression

Publicly available single-cell RNA-seq datasets from the posterior segment, anterior segment, and retina were used to plot the single-cell expression patterns of our prioritized genes across a variety of ocular cell types (*48–50*).

### Cell type-specific enrichment

To identify ocular cell types that are enriched for cell type-specific expression of genes that mapped to GWAS loci based on colocalizing e/sQTLs, we used a previously described pipeline, ECLIPSER (*34*). Briefly, ECLIPSER operates by determining if genes mapped to GWAS loci for a specific complex disease or trait based on e/sQTLs or other functional genomic data show greater cell type-specific expression compared to genes linked to a background set of GWAS loci from thousands of unrelated traits. The fundamental principle of ECLIPSER is that in a relevant tissue, multiple disease-associated genes will exhibit elevated expression in pathogenic cell types relative to non-pathogenic cells, to a greater extent than genes associated with unrelated traits.

The cell type-specific expression data we used was previously described in Hamel et. al, 2024 (*35*). It is single-nucleus RNA-seq of 13 tissues dissected from non-diseased human eyes, including central cornea, corneoscleral wedge (CSW), trabecular meshwork (TM) including Schlemm’s canal, iris, ciliary body (CB), lens (all from anterior segment(*51*)), peripheral and macular retina(*52, 53*), the optic nerve head (ONH), optic nerve (ON), peripapillary sclera (PPS), peripheral sclera, and choroid (all from posterior segment(*54*)). GWAS-specific genome-wide significant loci from the EUR and cross-ancestry meta-analysis were used as input for enrichment.

### List of **S**upplementary Materials

Fig. S1. Summary of colocalizing genes (e/sGenes) from the European meta-analysis results across GTEx tissues and retina.

Fig. S2. Cross-ancestry GWAS enrichment of specific cell types in ONH and anterior segment.

Fig. S3. EUR GWAS enrichment of specific cell types in ONH and anterior segment.

Fig. S4. Single-cell expression of genes in various cell types in anterior segment and optic nerve.

SI File 1. Summary statistics of LD-independent loci whose P values were lower than 1 x 10^-5^ for each endophenotype GWAS in the EUR discovery subset.

SI File 2. Summary statistics of LD-independent loci passing genome-wide significance in the EUR, AFR, ASN, and cross-ancestry meta-analyses.

SI File 3. Forest plots of genome-wide significant loci showing the consistency of effect sizes across different ancestry groups.

SI File 4. Genomic inflation factor statistics for all GWAS and meta-analyses.

SI File 5. Comparison of identified genome-wide significant loci with previously published associations for glaucoma and related traits.

SI File 6. MAGMA gene-based association tests results for the EUR, AFR, ASN, and cross-ancestry meta-analyses.

SI File 7. Complete results from the eCAVIAR colocalization analysis with EUR and cross-ancestry meta-analyses genome-wide significant loci with GTEx and retina QTLs.

SI File 8. ECLIPSER cell-type enrichment analysis results for the genome-wide significant loci from the EUR and cross-ancestry meta-analyses.

SI File 9. GSMR results for the significant genes after Bonferroni correction.

SI File 10. MAGMA gene-set enrichment analysis results of significant gene sets that passed Bonferroni correction.

## Data Availability

The GWAS summary statistics generated in this study are available in Figshare (10.6084/m9.figshare.30227764).

## Acknowledgments

We are grateful to the patients of the MEE glaucoma clinic and the participants of the UK Biobank whose contributions were essential for this research. We would also like to acknowledge Yuyang Luo, Puja Mehta, and Susan Xu who provided technical assistance in some of the computational tools.

## Funding

National Institutes of Health grants R01 EY036518 (NZ), R01 EY036222 (MW), R01 EY030575 (TE), NIH/NEI R01EY031424 (AVS)

P30EY003790 (TE), P30EY014104 (AVS)

Research to Prevent Blindness (NZ, TE)

## Author contributions

Conceptualization: NZ, AVS, JW, LC, SKH, TE, MW, ME, YZ

Methodology: NZ, AVS, JW, LC, SKH, TE, MW, ME

Investigation: NZ, AVS, JW, LC, SKH, TE, MW, ME, YZ

Visualization: LC, NZ, AVS

Funding acquisition: NZ, AVS, JW, TE, MW, ME

Project administration: NZ, AVS, JW, LC, SKH, TE, MW, ME, YZ

Supervision: NZ, AVS, JW, TE, MW, ME

Writing – original draft: LC

Writing – review & editing: LC, NZ, AVS, JW, YZ

## Data and materials availability

The GWAS summary statistics generated in this study are available in Figshare (10.6084/m9.figshare.30227764). UK Biobank data are available by request through the UK Biobank Access Management System https://www.ukbiobank.ac.uk/. The 1000 Genomes phase 3 data is available at https://www. internationalgenome.org/. The GTEx protected data are available through the database of Genotypes and Phenotypes (dbGaP) (accession no. phs000424.v8). The processed GTEx eQTL and sQTL summary statistics are available on the GTEx portal (https://gtexportal.org/home/datasets). The EyeGEx retina eQTL summary statistics is available at https://visiongenomics.org. The snRNA-seq data for the anterior segment and macula are available in Gene Expression Omnibus (GEO) accession number GSE199013, for the optic nerve head and posterior tissues in GSE236566, and for the retina in GSE226108. All the results from our analyses (e.g., GWAS, MAGMA, colocalization, Mendelian randomization, cell-type enrichment) can be found in the Supplementary Data.

## Supplementary Figures

**Fig. S1.**
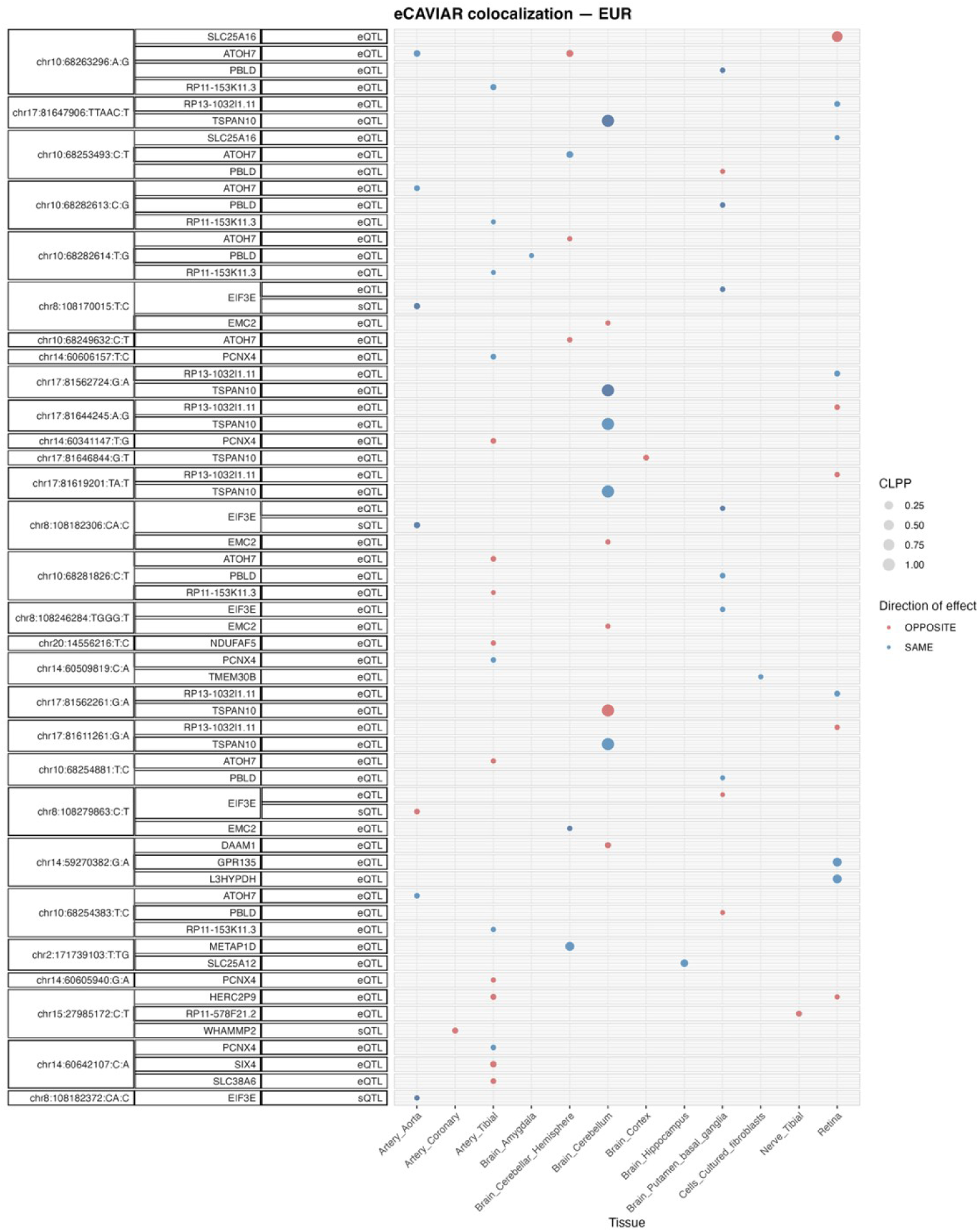
Colocalizing eQTLs and sQTLs in GTEx tissues and retina with genome-wide significant loci in EUR ML endophenotype meta-analyses. Genes with at least one significant colocalization result (CLPP>0.01) are shown for e/sQTLs tested across selected GTEx tissues and peripheral retina for the genome-wide significant loci from the EUR and the cross-ancestry endophenotype meta-analyses. Bubble size is proportional to the maximum colocalization posterior probability (CLPP) of all e/sVariants tested for the given gene, QTL type and tissue combination. Points are color-coded by the relative direction of effect between the QTL and the GWAS meta-analysis.

**Fig. S2.**
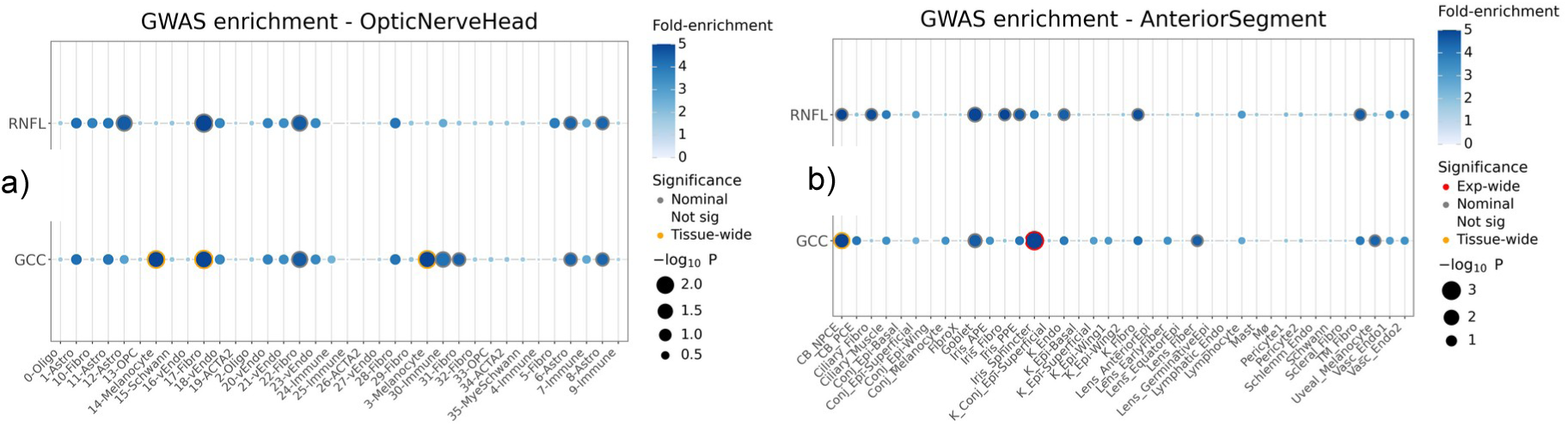
Cross-ancestry GWAS enrichment in specific cell types in a) optic nerve head, and b) anterior segment using ECLIPSER.

**Fig. S3.**
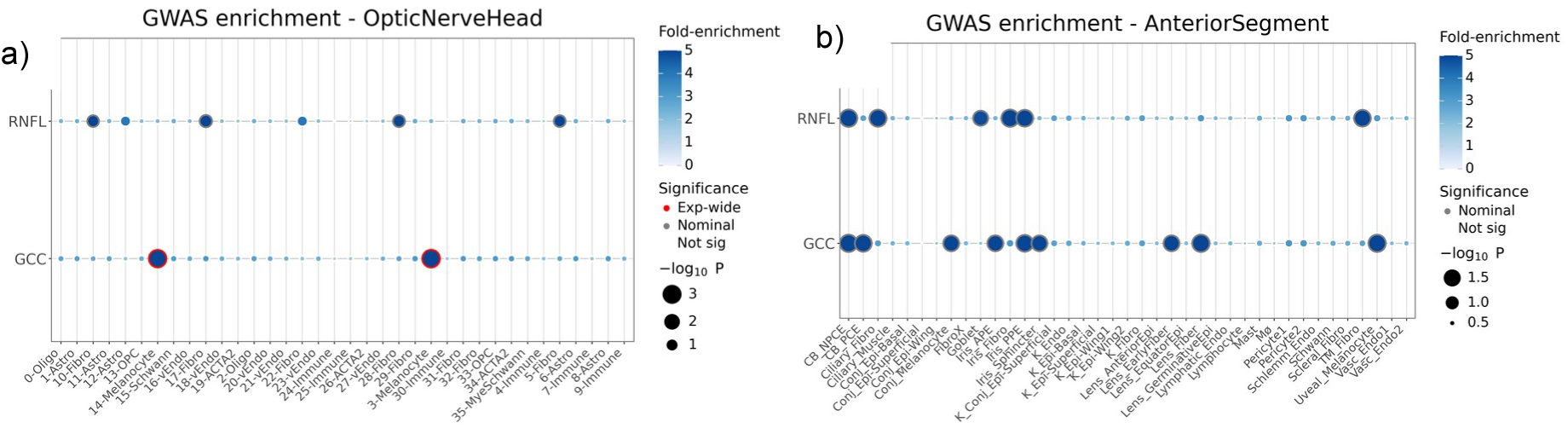
EUR GWAS enrichment in specific cell types in a) optic nerve head, and b) anterior segment using ECLIPSER.

**Fig. S4.**
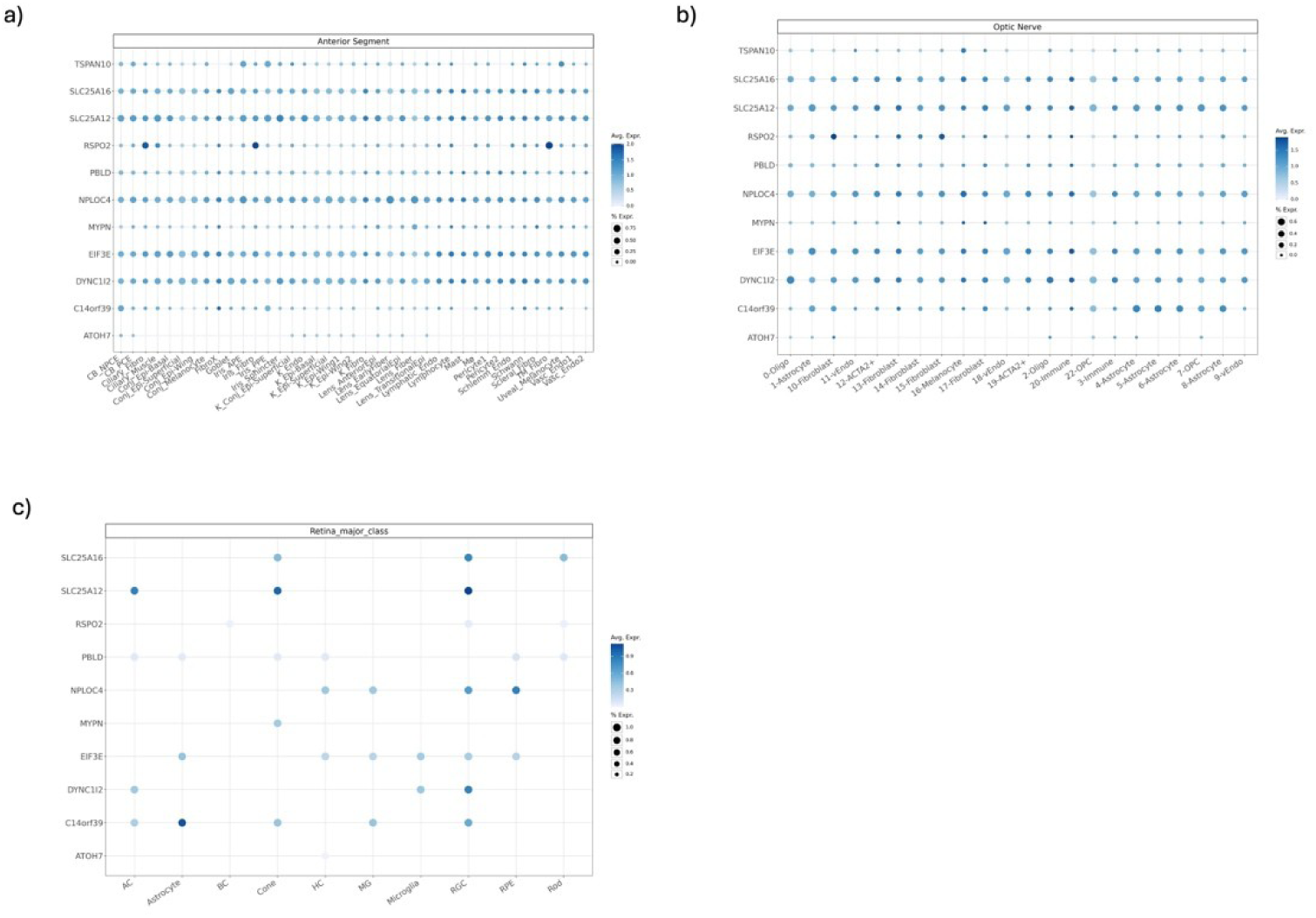
Single-cell expression of genes in various cell types in a) anterior segment, b) optic nerve head, optic nerve and surrounding posterior tissues, and c) retina. Single nucleus RNA-seq for the anterior segment was taken from van Zyl et al., 2022 (*48*) and for the optic nerve head and surrounding posterior tissues from Monavarfeshani, Yan, *et al.,* 2023 (*49*).

